# Salt Added to Food and Body Mass Index: A Bidirectional Mendelian Randomization Study

**DOI:** 10.1101/2020.06.02.20120097

**Authors:** Long Zhou, Xiaoxiao Wen, Liancheng Zhao, Yan Yu

## Abstract

**Background and objective:** Observational studies suggest that dietary sodium (salt) intake may be associated with body mass index (BMI). However, these findings may be biased by confounding and reverse causality. The present study aimed to apply a bidirectional Mendelian Randomization (MR) framework to determine the causal association between salt added to food (do not include salt used in cooking) and BMI by integrating summary-level genome-wide association study (GWAS) data.

**Methods:** We performed two-sample MR analyses using summary statistics of GWAS. Inverse-variance weighted (IVW) method was used to analyze the effect of the preference of salt added to food on BMI. We used maximum likelihood estimation and random effect model as auxiliary verification. A bidirectional MR analysis with BMI as the exposure and salt added to food as the outcome was also performed.

**Results:** We identified 74 single nucleotide polymorphisms (SNPs) that were genome-wide significant (*P* < 5×10^-8^) for the preference of salt added to food in the UK Biobank (n = 462,630) and were investigated for their association with BMI in a meta-analysis of 322,154 European-descent individuals from GWAS and Metabochip studies. The IVW method estimate indicated that the preference of salt added to food was positively associated with BMI (β = 0.1416, SE = 0.0576, *P* = 0.0139). Results from maximum likelihood estimation (β = 0.1476, SE = 0.0363, *P <* 0.0001) and the random effect model (β = 0.1411, SE = 0.0572, *P* = 0.0137) were consistent with the IVW. Bidirectional MR analyses suggested that BMI was not associated with the preference of salt added to food.

**Conclusion:** Our results provided qualitative evidence supporting a causal relationship between salt intake and BMI.

## Introduction

High sodium (salt) intake is one of the leading risk factors for disease burden across the world (1). Excess sodium intake has been identified as a major risk factor for high blood pressure (BP) (2,3) and thus leading to a higher risk of cardiovascular disease (CVD) (4,5). It was generally accepted that the relationship between high sodium intake and CVD was mediated by elevated BP or hypertension because high BP or hypertension is a well-established risk factor for CVD, including stroke and coronary heart disease (6,7). In addition to elevating BP, whether high sodium intake can increase the risk of CVD through other ways is a worthwhile scientific issue. In the past few years, several epidemiological studies have found that dietary sodium intake may be positively associated with body mass index (BMI) as well as the odds of overweight or obesity (8-10). The International Study of Macro-/Micro-nutrients and Blood Pressure (INTERMAP Study) showed that dietary salt intake 1 g/d higher was significantly associated with BMI higher by 0.28 in Japan, 0.10 in China, 0.42 in the United Kingdom, and 0.52 in the United States after adjustment for physical activity and energy intake (10). These findings suggest that overweight/obesity may also serve as a mediator between high sodium intake and the risk of CVD because overweight/obesity is also a well-known risk factor for CVD (1,11,12). To date, studies on the relationship between dietary sodium intake and BMI, as well as overweight/obesity, were almost cross-sectional designs, which unable to give a causal relationship. In the absence of high-quality randomized clinical trial (RCT) data, the mendelian randomization (MR) can be applied to strengthen or refute the causality of biomarkers in disease etiology (13). We applied a two-sample MR framework to determine the causal association between the preference of salt added to food (do not include salt used in cooking) and BMI by integrating summary-level genome-wide association study (GWAS) data.

## Methods

### Data sources and single nucleotide polymorphism (SNP) selection

Effect estimates for SNPs associated with the preference of salt added to food were obtained from UK Biobank, which is the largest GWAS in 462,630 European-ancestry individuals. Food preferences are the main reason for choosing to eat what we do. Understanding the biological and environmental factors influence food preferences and how they are related to food intake and health outcomes may help to improve both dietary guidelines and interventions to help people embrace a healthier lifestyle. UK Biobank provides a unique opportunity to understand food preferences better. Salt added to food in UK Biobank includes 4 levels: Never/rarely, Sometimes, Usually, and Always. Previous studies have shown that the preference of adding salt to cooked food could reflect the relative level of salt intake to a certain extent (14-16). We selected SNPs associated with the preference for salt added to food at a genome-wide significant level (*P* < 5×10^-8^). Effect estimates of these salt-associated SNPs on the BMI levels were assessed using the summary statistics from a meta-analysis of 322,154 European-descent individuals from GWAS and Metabochip studies (17). All GWASs have existing ethical permissions from their respective institutional review boards and include participant informed consent.

### SNP validation

One requirement of MR is that the selected SNPs must not be in Linkage disequilibrium (LD) with each other, since if a selected SNP is highly correlated with other risk factor loci, this may result in confounding (13). In our analysis, SNPs were removed if their measured LD had an r^2^ greater than 0.001. We retained the SNP most strongly associated with the preference of salt added to food by p-value.

The SNPs selected for MR analysis cannot be directly related to the outcome. In order to meet this requirement, we removed those SNPs that are directly related (*P* < 5×10^-8^, a conventional threshold for declaring statistical significance in GWAS) to BMI.

An essential assumption of MR is that each SNP must only influence the risk of the outcome through the exposure, as the inclusion of SNPs that contribute through a pleiotropic pathway could bias estimates. We used MR-Egger regression to assess for the presence of pleiotropy (18). Besides, to ensure that there is no correlation between SNPs and confounding factors, we searched these selected SNPs in the PubMed database. Those SNPs related to possible confounding factors were removed from MR analysis.

### Statistical analysis

The summary-level GWAS data for the exposure and outcome were computed from two independent studies with individual-level SNP genotypes. We harmonized the summary data for salt added to food and BMI so that the effect estimates of SNPs on exposure and outcome were shown for the same alleles. We mainly used the inverse-variance weighted (IVW) method to estimate the effect of the preference of salt added to food on BMI (19,20). To improve the reliability of the results, we also used maximum likelihood estimation and random effect model as auxiliary verification. To ensure that the inclusion of SNPs did not introduce random error into our results, we conducted sensitivity analyses using the leave-one-out method. In addition, we performed a bidirectional MR analysis with BMI as the exposure and salt added to food as the outcome to explore whether BMI influences the preference of salt added to food. All analyses were performed using R version 3.6.3 (R Foundation for Statistical Computing, Vienna, Austria).

## Results

After screening and validation, we identified 74 SNPs that were significantly associated with the preference of salt added to food (*P* < 5×10^-8^) and independent of BMI (*P* > 5×10^-8^). The characteristics of the selected SNPs are presented in **Table 1**.

**Table 1.**
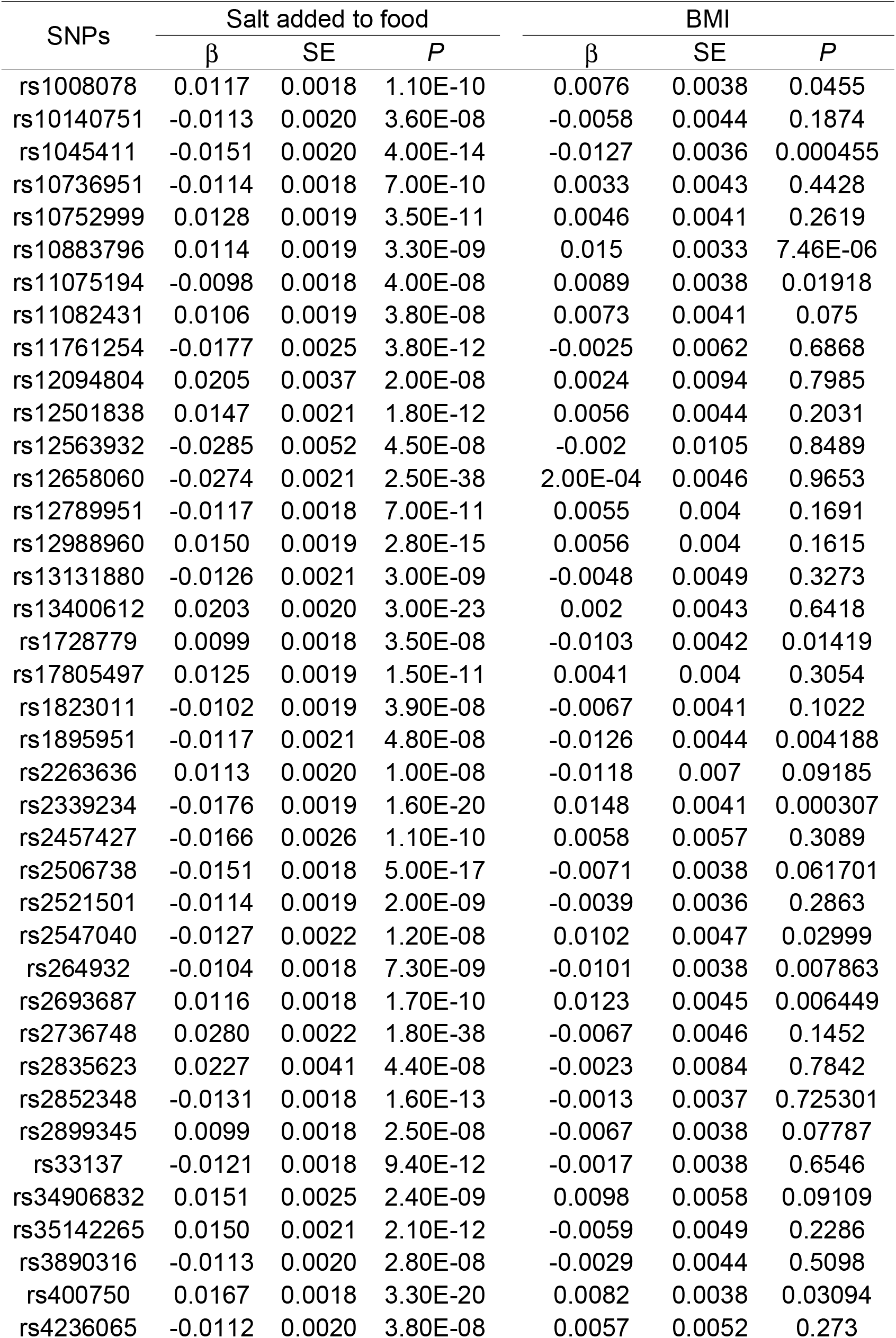
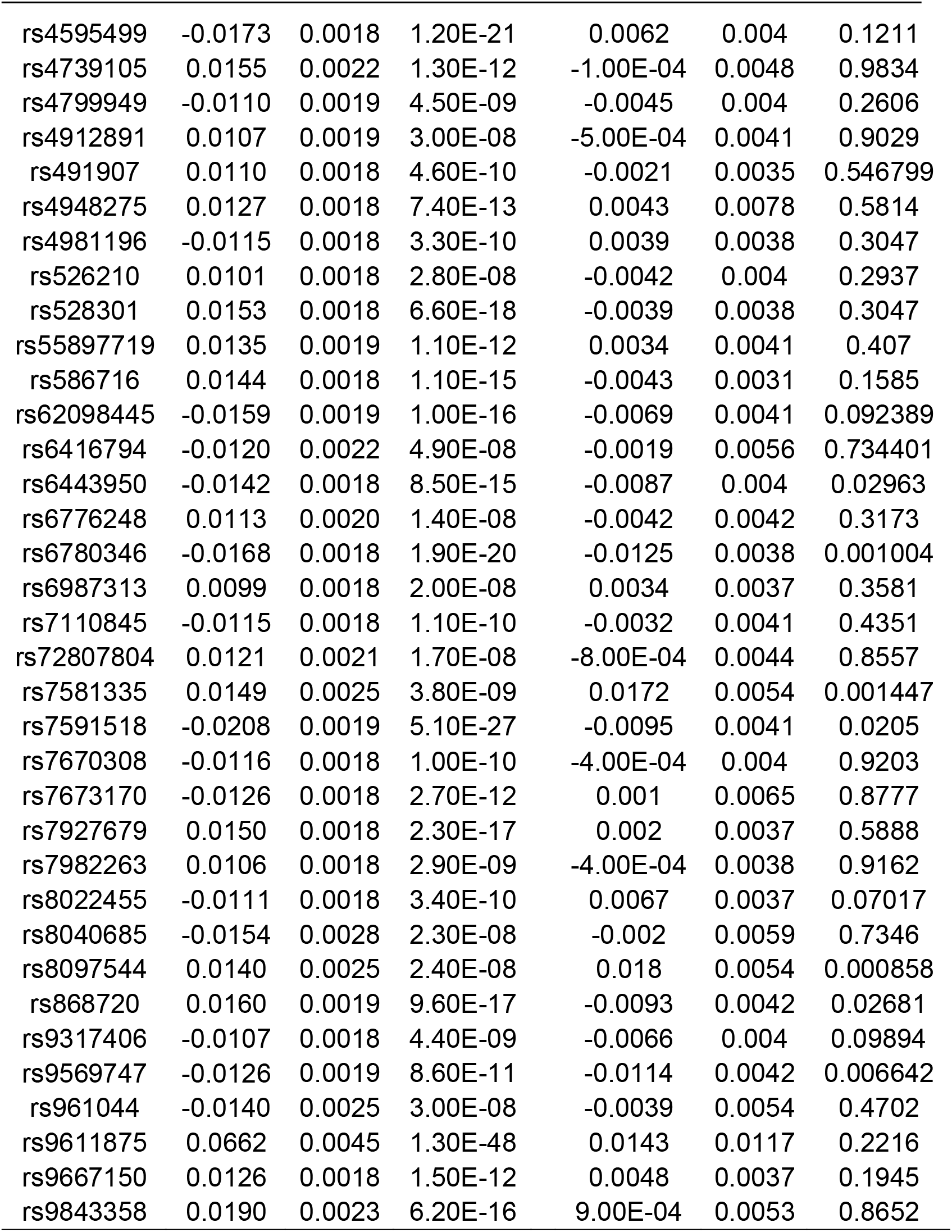
Characteristics of the selected single-nucleotide polymorphisms associated with salt added to food and their associations with body mass index

| SNPs | Salt added to food |  |  | BMI |  |  |
| --- | --- | --- | --- | --- | --- | --- |
| | $\beta$ | SE | *P* | $\beta$ | SE | *P* |
| rs1008078 | 0.0117 | 0.0018 | 1.10E-10 | 0.0076 | 0.0038 | 0.0455 |
| rs10140751 | -0.0113 | 0.0020 | 3.60E-08 | -0.0058 | 0.0044 | 0.1874 |
| rs1045411 | -0.0151 | 0.0020 | 4.00E-14 | -0.0127 | 0.0036 | 0.000455 |
| rs10736951 | -0.0114 | 0.0018 | 7.00E-10 | 0.0033 | 0.0043 | 0.4428 |
| rs10752999 | 0.0128 | 0.0019 | 3.50E-11 | 0.0046 | 0.0041 | 0.2619 |
| rs10883796 | 0.0114 | 0.0019 | 3.30E-09 | 0.015 | 0.0033 | 7.46E-06 |
| rs11075194 | -0.0098 | 0.0018 | 4.00E-08 | 0.0089 | 0.0038 | 0.01918 |
| rs11082431 | 0.0106 | 0.0019 | 3.80E-08 | 0.0073 | 0.0041 | 0.075 |
| rs11761254 | -0.0177 | 0.0025 | 3.80E-12 | -0.0025 | 0.0062 | 0.6868 |
| rs12094804 | 0.0205 | 0.0037 | 2.00E-08 | 0.0024 | 0.0094 | 0.7985 |
| rs12501838 | 0.0147 | 0.0021 | 1.80E-12 | 0.0056 | 0.0044 | 0.2031 |
| rs12563932 | -0.0285 | 0.0052 | 4.50E-08 | -0.002 | 0.0105 | 0.8489 |
| rs12658060 | -0.0274 | 0.0021 | 2.50E-38 | 2.00E-04 | 0.0046 | 0.9653 |
| rs12789951 | -0.0117 | 0.0018 | 7.00E-11 | 0.0055 | 0.004 | 0.1691 |
| rs12988960 | 0.0150 | 0.0019 | 2.80E-15 | 0.0056 | 0.004 | 0.1615 |
| rs13131880 | -0.0126 | 0.0021 | 3.00E-09 | -0.0048 | 0.0049 | 0.3273 |
| rs13400612 | 0.0203 | 0.0020 | 3.00E-23 | 0.002 | 0.0043 | 0.6418 |
| rs1728779 | 0.0099 | 0.0018 | 3.50E-08 | -0.0103 | 0.0042 | 0.01419 |
| rs17805497 | 0.0125 | 0.0019 | 1.50E-11 | 0.0041 | 0.004 | 0.3054 |
| rs1823011 | -0.0102 | 0.0019 | 3.90E-08 | -0.0067 | 0.0041 | 0.1022 |
| rs1895951 | -0.0117 | 0.0021 | 4.80E-08 | -0.0126 | 0.0044 | 0.004188 |
| rs2263636 | 0.0113 | 0.0020 | 1.00E-08 | -0.0118 | 0.007 | 0.09185 |
| rs2339234 | -0.0176 | 0.0019 | 1.60E-20 | 0.0148 | 0.0041 | 0.000307 |
| rs2457427 | -0.0166 | 0.0026 | 1.10E-10 | 0.0058 | 0.0057 | 0.3089 |
| rs2506738 | -0.0151 | 0.0018 | 5.00E-17 | -0.0071 | 0.0038 | 0.061701 |
| rs2521501 | -0.0114 | 0.0019 | 2.00E-09 | -0.0039 | 0.0036 | 0.2863 |
| rs2547040 | -0.0127 | 0.0022 | 1.20E-08 | 0.0102 | 0.0047 | 0.02999 |
| rs264932 | -0.0104 | 0.0018 | 7.30E-09 | -0.0101 | 0.0038 | 0.007863 |
| rs2693687 | 0.0116 | 0.0018 | 1.70E-10 | 0.0123 | 0.0045 | 0.006449 |
| rs2736748 | 0.0280 | 0.0022 | 1.80E-38 | -0.0067 | 0.0046 | 0.1452 |
| rs2835623 | 0.0227 | 0.0041 | 4.40E-08 | -0.0023 | 0.0084 | 0.7842 |
| rs2852348 | -0.0131 | 0.0018 | 1.60E-13 | -0.0013 | 0.0037 | 0.725301 |
| rs2899345 | 0.0099 | 0.0018 | 2.50E-08 | -0.0067 | 0.0038 | 0.07787 |
| rs33137 | -0.0121 | 0.0018 | 9.40E-12 | -0.0017 | 0.0038 | 0.6546 |
| rs34906832 | 0.0151 | 0.0025 | 2.40E-09 | 0.0098 | 0.0058 | 0.09109 |
| rs35142265 | 0.0150 | 0.0021 | 2.10E-12 | -0.0059 | 0.0049 | 0.2286 |
| rs3890316 | -0.0113 | 0.0020 | 2.80E-08 | -0.0029 | 0.0044 | 0.5098 |
| rs400750 | 0.0167 | 0.0018 | 3.30E-20 | 0.0082 | 0.0038 | 0.03094 |
| rs4236065 | -0.0112 | 0.0020 | 3.80E-08 | 0.0057 | 0.0052 | 0.273 |
| rs4595499 | -0.0173 | 0.0018 | 1.20E-21 | 0.0062 | 0.004 | 0.1211 |
| rs4739105 | 0.0155 | 0.0022 | 1.30E-12 | -1.00E-04 | 0.0048 | 0.9834 |
| rs4799949 | -0.0110 | 0.0019 | 4.50E-09 | -0.0045 | 0.004 | 0.2606 |
| rs4912891 | 0.0107 | 0.0019 | 3.00E-08 | -5.00E-04 | 0.0041 | 0.9029 |
| rs491907 | 0.0110 | 0.0018 | 4.60E-10 | -0.0021 | 0.0035 | 0.546799 |
| rs4948275 | 0.0127 | 0.0018 | 7.40E-13 | 0.0043 | 0.0078 | 0.5814 |
| rs4981196 | -0.0115 | 0.0018 | 3.30E-10 | 0.0039 | 0.0038 | 0.3047 |
| rs526210 | 0.0101 | 0.0018 | 2.80E-08 | -0.0042 | 0.004 | 0.2937 |
| rs528301 | 0.0153 | 0.0018 | 6.60E-18 | -0.0039 | 0.0038 | 0.3047 |
| rs55897719 | 0.0135 | 0.0019 | 1.10E-12 | 0.0034 | 0.0041 | 0.407 |
| rs586716 | 0.0144 | 0.0018 | 1.10E-15 | -0.0043 | 0.0031 | 0.1585 |
| rs62098445 | -0.0159 | 0.0019 | 1.00E-16 | -0.0069 | 0.0041 | 0.092389 |
| rs6416794 | -0.0120 | 0.0022 | 4.90E-08 | -0.0019 | 0.0056 | 0.734401 |
| rs6443950 | -0.0142 | 0.0018 | 8.50E-15 | -0.0087 | 0.004 | 0.02963 |
| rs6776248 | 0.0113 | 0.0020 | 1.40E-08 | -0.0042 | 0.0042 | 0.3173 |
| rs6780346 | -0.0168 | 0.0018 | 1.90E-20 | -0.0125 | 0.0038 | 0.001004 |
| rs6987313 | 0.0099 | 0.0018 | 2.00E-08 | 0.0034 | 0.0037 | 0.3581 |
| rs7110845 | -0.0115 | 0.0018 | 1.10E-10 | -0.0032 | 0.0041 | 0.4351 |
| rs72807804 | 0.0121 | 0.0021 | 1.70E-08 | -8.00E-04 | 0.0044 | 0.8557 |
| rs7581335 | 0.0149 | 0.0025 | 3.80E-09 | 0.0172 | 0.0054 | 0.001447 |
| rs7591518 | -0.0208 | 0.0019 | 5.10E-27 | -0.0095 | 0.0041 | 0.0205 |
| rs7670308 | -0.0116 | 0.0018 | 1.00E-10 | -4.00E-04 | 0.004 | 0.9203 |
| rs7673170 | -0.0126 | 0.0018 | 2.70E-12 | 0.001 | 0.0065 | 0.8777 |
| rs7927679 | 0.0150 | 0.0018 | 2.30E-17 | 0.002 | 0.0037 | 0.5888 |
| rs7982263 | 0.0106 | 0.0018 | 2.90E-09 | -4.00E-04 | 0.0038 | 0.9162 |
| rs8022455 | -0.0111 | 0.0018 | 3.40E-10 | 0.0067 | 0.0037 | 0.07017 |
| rs8040685 | -0.0154 | 0.0028 | 2.30E-08 | -0.002 | 0.0059 | 0.7346 |
| rs8097544 | 0.0140 | 0.0025 | 2.40E-08 | 0.018 | 0.0054 | 0.000858 |
| rs868720 | 0.0160 | 0.0019 | 9.60E-17 | -0.0093 | 0.0042 | 0.02681 |
| rs9317406 | -0.0107 | 0.0018 | 4.40E-09 | -0.0066 | 0.004 | 0.09894 |
| rs9569747 | -0.0126 | 0.0019 | 8.60E-11 | -0.0114 | 0.0042 | 0.006642 |
| rs961044 | -0.0140 | 0.0025 | 3.00E-08 | -0.0039 | 0.0054 | 0.4702 |
| rs9611875 | 0.0662 | 0.0045 | 1.30E-48 | 0.0143 | 0.0117 | 0.2216 |
| rs9667150 | 0.0126 | 0.0018 | 1.50E-12 | 0.0048 | 0.0037 | 0.1945 |
| rs9843358 | 0.0190 | 0.0023 | 6.20E-16 | 9.00E-04 | 0.0053 | 0.8652 |

We used MR-Egger regression to assess for the presence of pleiotropy, and the result showed that the intercept term was centered at the origin with a confidence interval including the null (β_0_=0.0014, 95% CI −0.0041, 0.0069, *P* = 0.6121), suggesting that pleiotropy had not unduly influenced the results.

The regression lines obtained from IVW, maximum likelihood estimation, and the random effect model were basically in coincidence, which suggests that the results would not be influenced by different analytic methods (**Figure 1**). In addition, we inspected the funnel plot for any asymmetry, which would be suggestive of pleiotropy, and found the plot to the symmetrical (**Figure 2**).

**Figure 1.**
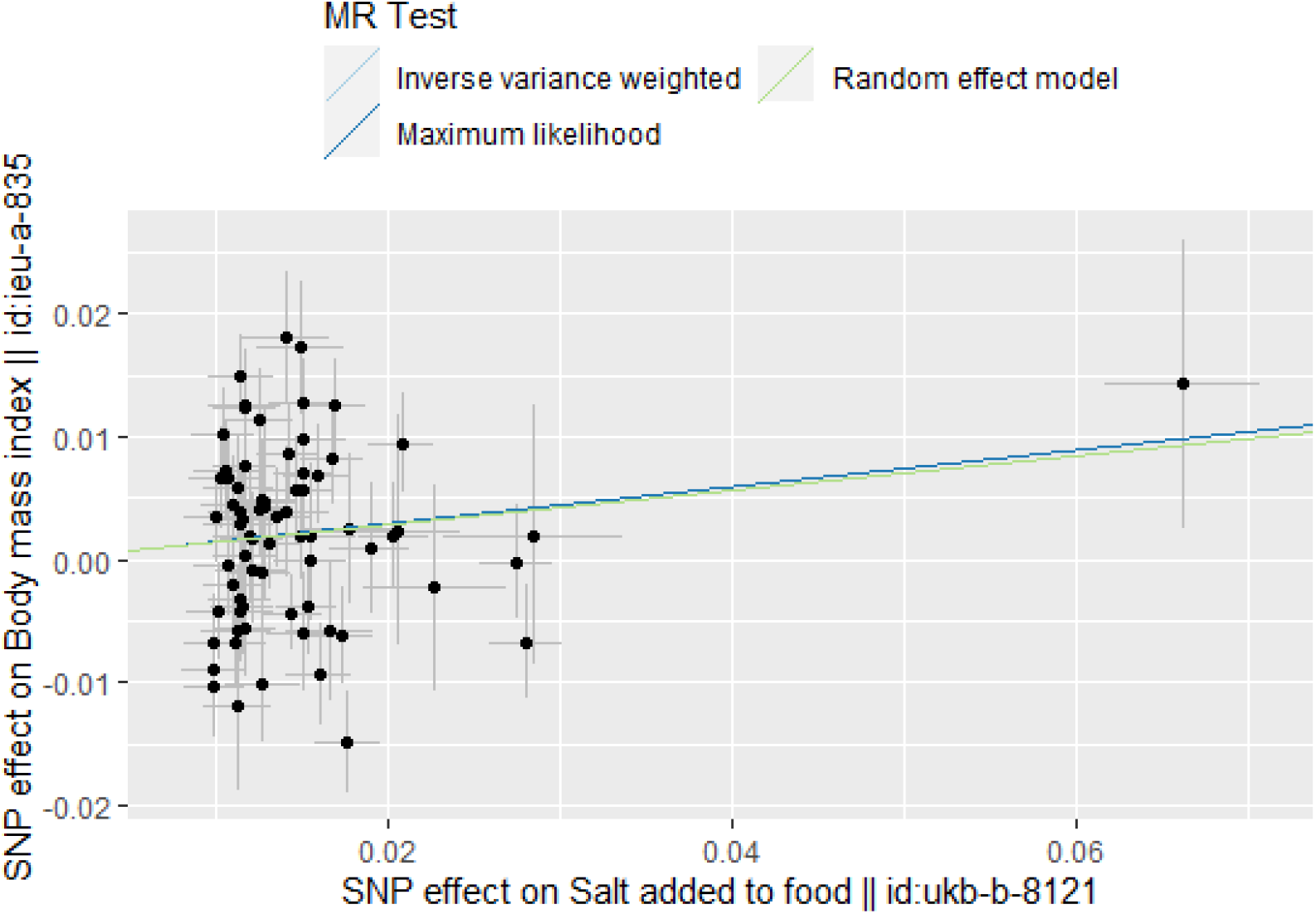
Scatter plot for the preference of salt added to food on BMI analysis

**Figure 2.**
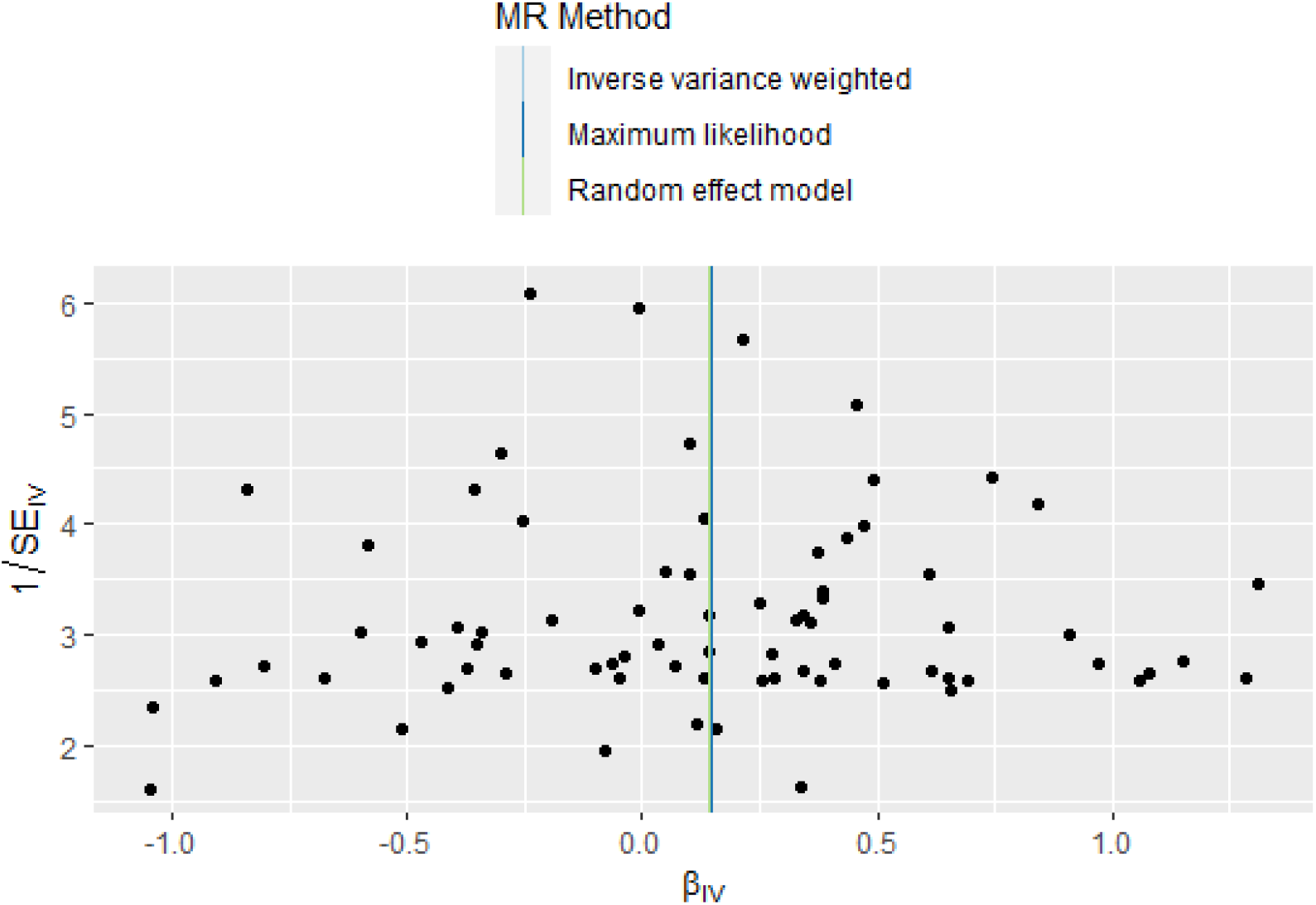
Funnel plot for the preference of salt added to food on BMI analysis

The IVW method estimate indicated that the preference of salt added to food was positively associated with BMI (β = 0.1416, SE = 0.0576, *P* = 0.0139). Results from maximum likelihood estimation (β = 0.1476, SE = 0.0363, *P* < 0.0001) and the random effect model (β = 0.1411, SE = 0.0572, *P* = 0.0137) were consistent with the IVW. The forest plot of each SNP estimates and pooled results using different methods are presented in **Figure 3**. MR leave-one-out sensitivity analysis showed that the pooled effect of the preference of salt added to food on BMI was significant (β = 0.1416, SE = 0.0576, *P* = 0.0139), which suggests a robust relationship between the preference of salt added to food and BMI (**Figure 4**).

**Figure 3.**
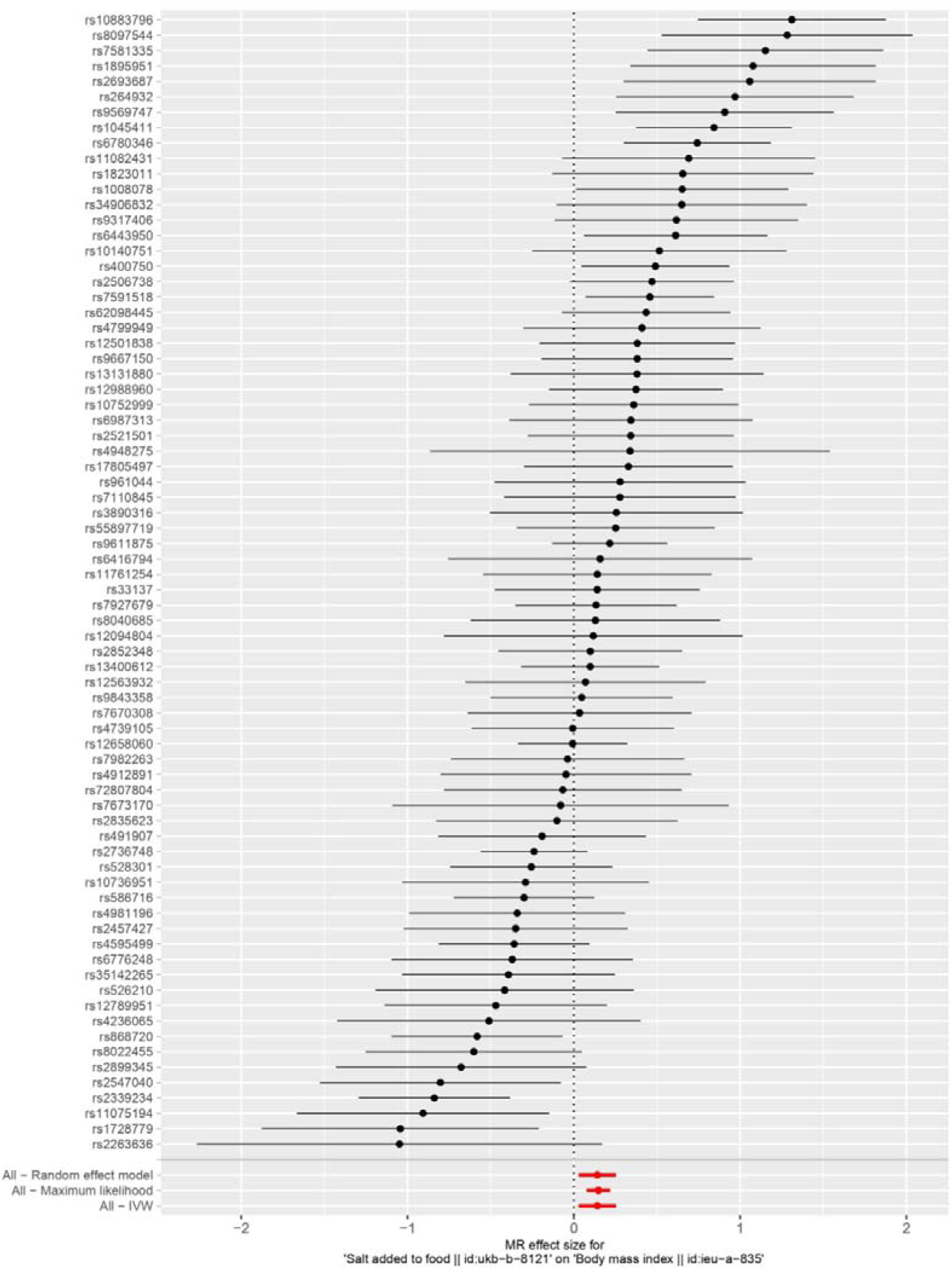
Forest plot of each SNP estimates and pooled results using different methods

**Figure 4.**
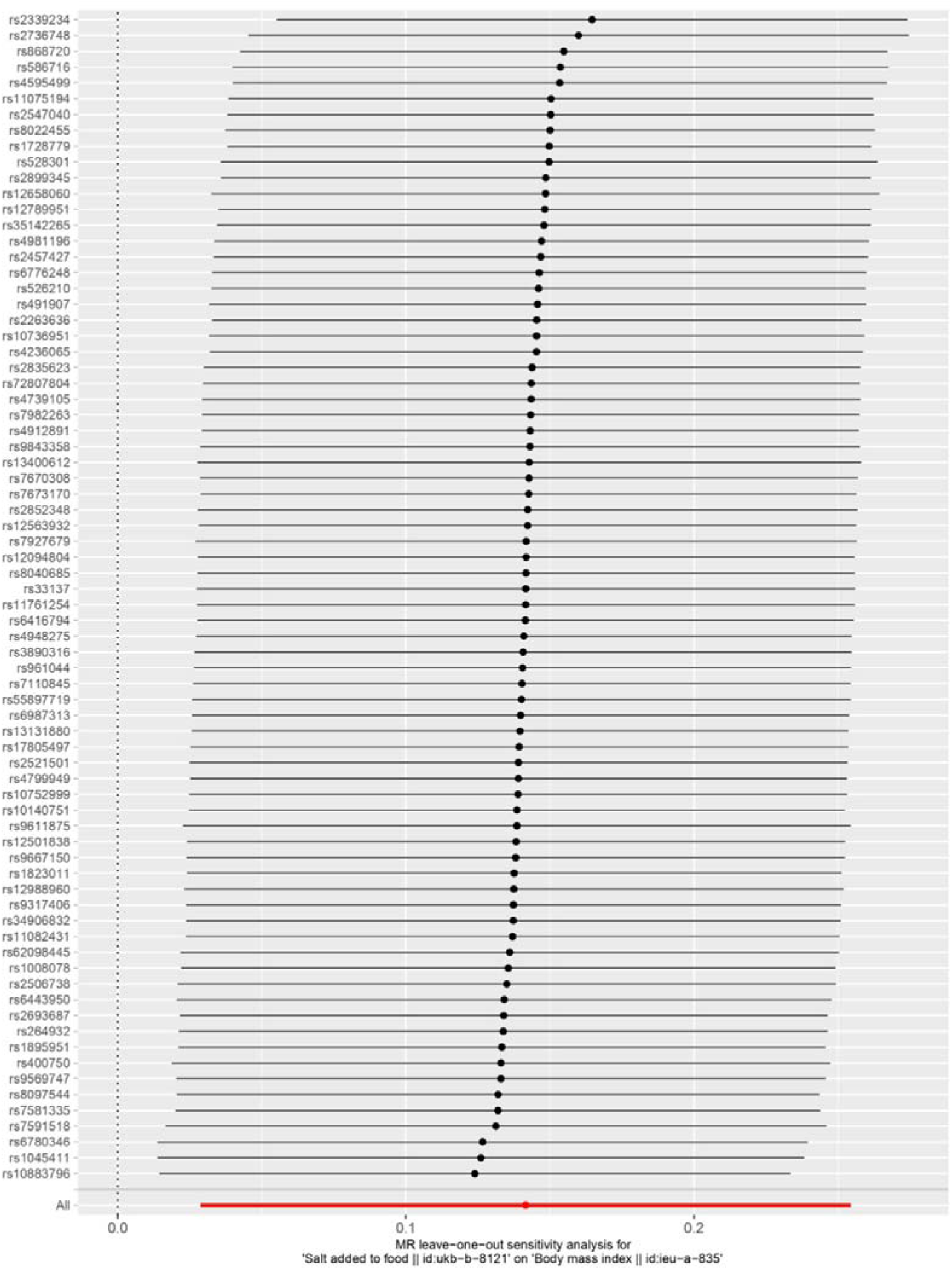
Forest plot for MR leave-one-out sensitivity analysis

To further clarify the causal relationship between the preference of salt added to food and BMI and exclude the possible impact of a reverse causal relationship, we further performed a bidirectional MR analysis with BMI as the exposure and the preference of salt added to food as the outcome. We identified 67 SNPs that were significantly associated with BMI (*P* < 5×10^-8^) and independent of the preference of salt added to food (*P* > 5×10^-8^). There was no significant pleiotropy (β_0_=0.0008, 95% CI −0.0022, 0.0038, *P* = 0.5909). MR analyses showed that BMI was not associated with the preference of salt added to food (**Figure 5**).

**Figure 5.**
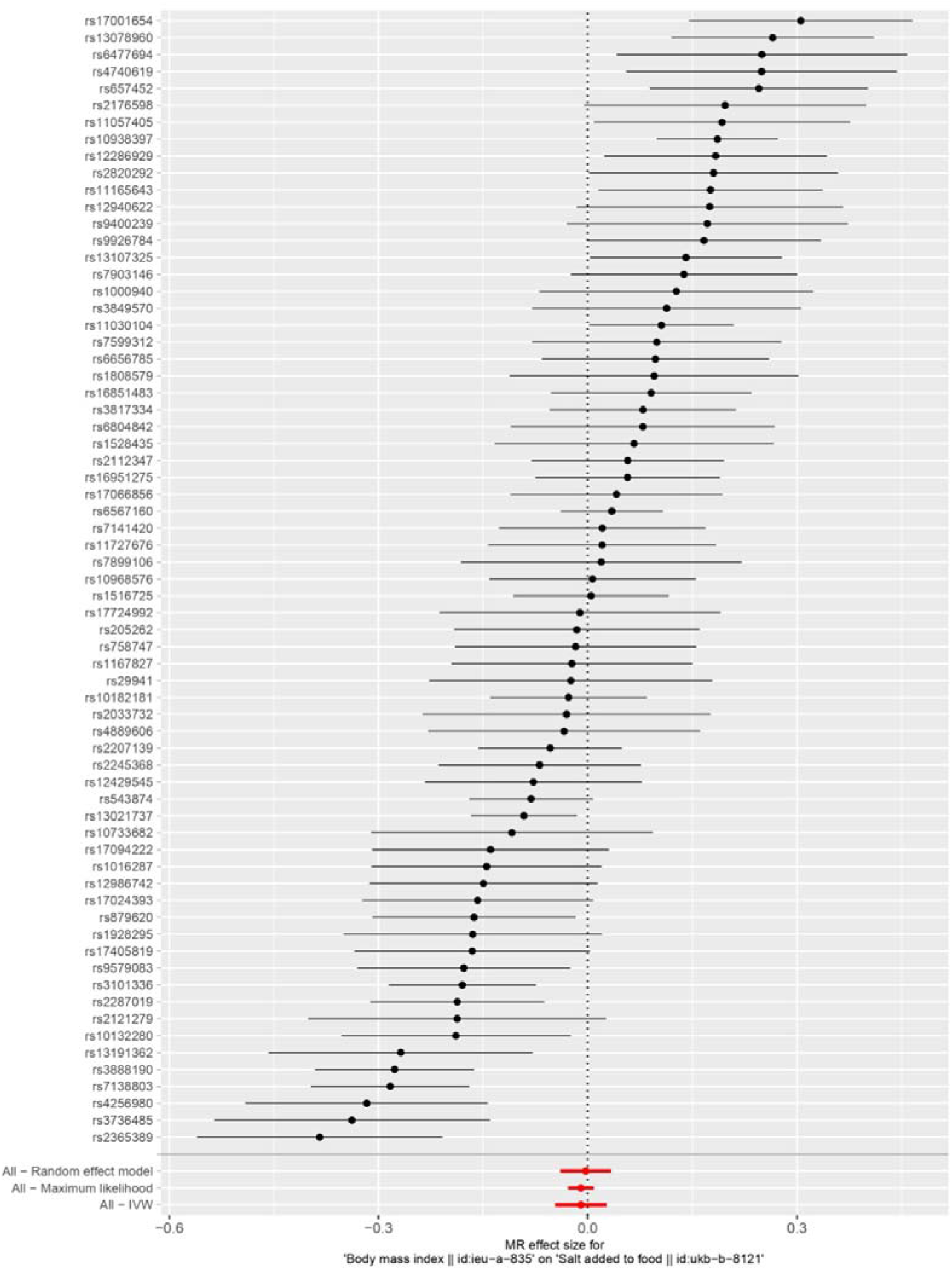
Forest plot for bidirectional MR analysis with BMI as the exposure and the preference of salt added to food as the outcome

## Discussion

Using a bidirectional MR study design, based on the largest GWAS summary statistics available on the preference of salt added to food and BMI, our analyses showed a positive association between the preference of salt added to food and BMI, and this relationship was causal. These findings have important public health implications in that reducing salt intake, in addition to lowering BP, may also lower BMI, and thereby reduce the risk of CVD.

The prevalence of obesity has increased dramatically worldwide over the last decades (12). A systematic estimation showed that 57.8% of the world population would be overweight or obese by the year 2030 if the current trends continue (21). Numerous studies have demonstrated that obesity is an important risk factor for CVD. The Manitoba Study prospectively investigated 3,983 men for 26 years and found that a high BMI was significantly associated with the development of myocardial infarction (MI), sudden death and coronary insufficiency, or suspected MI (22). The Framingham Heart Study showed that overweight/obesity (BMI ≥ 25 kg/m^2^) was associated with a 23% higher risk of coronary heart disease for men and 15% for women (23). The Global Burden of Metabolic Risk Factors for Chronic Disease Collaboration conducted a meta-analysis on 97 cohort studies, with 1.8 million participants reported significant BMI effects on the development of CHD and stroke. The Hazard Ratio for each 5 kg/m^2^ higher BMI was 1.27 (95% CI 1.23-1.31) for coronary heart disease and 1.18 (1.14-1.22) for stroke after adjustment for confounders (24).

In the last few years, several studies have linked dietary salt intake with BMI in the general population. Although some of those previous studies had methodological problems, namely the use of either spot urine (25) or dietary recall methods (26,27) to estimate individual salt intake, which has been criticized for large bias in estimating individual salt intake (28,29), most of the studies reported a positive association between dietary salt intake and BMI (8-10,26,27,30-33). The INTERMAP Study is the largest international multicenter study on the relationship between salt intake and BMI to date. It used 2 timed 24-h urine collections to estimate salt intake and 4 in-depth multipass 24-h dietary recalls for estimating daily energy intake. After adjustment for several potential confounding factors including daily total energy intake and physical activity, salt intake 1 g/d higher was associated with BMI 0.28 (95% CI, 0.23, 0.34) higher in Japan, 0.10 (95% CI, 0.05, 0.14) higher in China, 0.42 (95% CI, 0.27, 0.56) higher in the United Kingdom, and 0.52 (95% CI, 0.45, 0.59) higher in the United States (10). However, almost all these studies were cross-sectional design, which cannot avoid inherent limitations such as unknown confounding factors and reverse causal association. In the present study, a bidirectional MR study was applied to identify the causal association between the preference of salt added to food and BMI. Results from the IVW method, maximum likelihood estimation, and the random effect model support that the genetically predicted the preference of salt added to food was positively and casually associated with BMI, which was consistent with the previous observational studies.

Our present study had several limitations. First, we used the preference of salt added to food as the primary exposure rather than the gold standard for estimating daily salt intake because there is no GWAS study of 24-hour urinary sodium so far. Nevertheless, the preference of adding salt to cooked food could reflect the relative level of long-term salt intake to a certain extent and is better than using a spot urine sample to estimate 24-hour sodium excretion. Second, the preference of salt added to food was based on self-reported answers in the UK Biobank, and different people could differently interpret the question. Third, it is impossible to quantitatively evaluate the relationship between salt intake and BMI when using the preference of salt added to food as the exposure.

In conclusion, our analysis provided qualitative evidence supporting a causal relationship between salt intake and BMI. Further studies using genetic variants of 24-hour urinary sodium excretion as exposure are needed to validate this finding.

## Data Availability

All data was publicly available through https://gwas.mrcieu.ac.uk/

